# Not in Education Employment or Training (NEET) Trajectories Across Adolescence and Early Adulthood and Subsequent Mental Health

**DOI:** 10.64898/2026.09.14.26362986

**Authors:** Alex S. F. Kwong, Esme Elsden

## Abstract

In 3,792 participants from the Avon Longitudinal Study of Parents and Children, we examined whether repeatedly being Not in Education, Employment or Training (NEET) between ages 16 and 33 was associated with mental health at age 33. Each additional NEET occasion was associated with higher odds of depression (OR: 1.29, 95%CI: 1.18–1.40), anxiety (1.21, 1.11– 1.32), lower life satisfaction (1.53, 1.40–1.68) and a recent suicide attempt (1.51, 1.28–1.77). Associations were dose-dependent for depression, life satisfaction and a recent suicide attempt, and strongest for first NEET at ages 25–33. Preventing repeated NEET and addressing mental health need at first NEET entry are potential intervention targets.

## Introduction

Not being in education, employment, or training (NEET) is associated with poorer mental, physical health and reduced long-term economic prospects [1,2]. NEET affects, on average, 14% of young people aged 18-24 years across high-income countries in the OECD [3]. The UK is an outlier in trajectory rather than level, as numbers exceeded one million in early 2026 [4], and the UK moved from the European average to among the highest rates within the last decade [3]. The composition has changed with a sharp rise in young people reporting work-limiting health conditions, making economic inactivity an urgent health concern [5].

Most previous research has measured NEET as a static factor, despite variation in its onset, duration and recurrence [1]. These is a need to understand how repeated occasions of NEET across adolescence and adulthood carry cumulative risk for later mental health, whether the age at which NEET first occurs matters, and whether sustained and recurrent patterns of NEET differ in their associations with subsequent mental health outcomes.

Longitudinal studies with repeated assessments offer an important opportunity to address these questions. Using data from a large UK population-based cohort, we examined whether the number, timing and type of NEET occasions across adolescence and early adulthood were associated with later mental health, adjusting for baseline mental health and a range of sociodemographic and family factors.

## Methods

Data were from the Avon Longitudinal Study of Parents and Children (ALSPAC), a population-based cohort that recruited pregnant women residing in Avon (South-West England) with expected delivery dates between 1 April 1991 and 31 December 1992; the cohort comprises 14,901 children, now aged 33-35 [6, 7]. Ethical approval was obtained from the ALSPAC Ethics and Law Committee and Local Research Ethics Committees. Full sample, measure and consent details are provided in the Supplement.

NEET status was prospectively self-reported on 11 assessments between ages 16 and 33 years. Four exposures were derived: a count of NEET occasions (0-11 occasions); an ordinal category (0, 1, 2, 3+ occasions); age at first NEET (never, 16-20, 21-24, 25-33 years); and type of NEET (never, one occasion only, sustained NEET [across 2 or more consecutive assessments], or recurrent NEET across non-consecutive assessments).

Outcomes at age 33 were depression (Short Mood and Feelings Questionnaire [SMFQ] [8]), anxiety (Generalized Anxiety Disorder Assessment-7 [GAD-7] [9]), lower life satisfaction (Satisfaction with Life Scale [SWLS] [10]), a recent suicide attempt within the last two years, dichotomised using established cut-offs.

We used logistic regression to explore associations between each NEET exposure and mental health outcomes, adjusted for sex, ethnicity, maternal education, social class, financial problems and postnatal depression at birth, number of childhood traumas, childhood academic attainment (Key Stage 2 scores), the corresponding mental health measure assessed at baseline, and the number of completed NEET assessments. Missing covariate data were handled using multiple imputation by chained equations [11]. *P* values were corrected across all tests using the Benjamini-Hochberg false discovery rate. Analyses were completed in Stata 18. Further methodological information can be found in the supplement.

## Results

A total of 3,792 individuals had complete data on NEET trajectories and mental health outcomes at age 33.

Each additional NEET occasion between ages 16 and 33 was associated with poorer mental health at age 33 (Figure 1; Supplementary Table 1), including higher odds of depression (OR: 1.29 [95%CI: 1.18, 1.40], *P*_*FDR*_=3.04×10^-08^), anxiety (1.21 [1.11, 1.32], *P*_*FDR*_=7.54×10^-05^), lower life satisfaction (1.53 [1.40, 1.68], *P*_*FDR*_=2.26×10^-19^) and a recent suicide attempt (1.51 [1.28, 1.77], *P*_*FDR*_=1.17×10^-06^).

**Figure 1.**
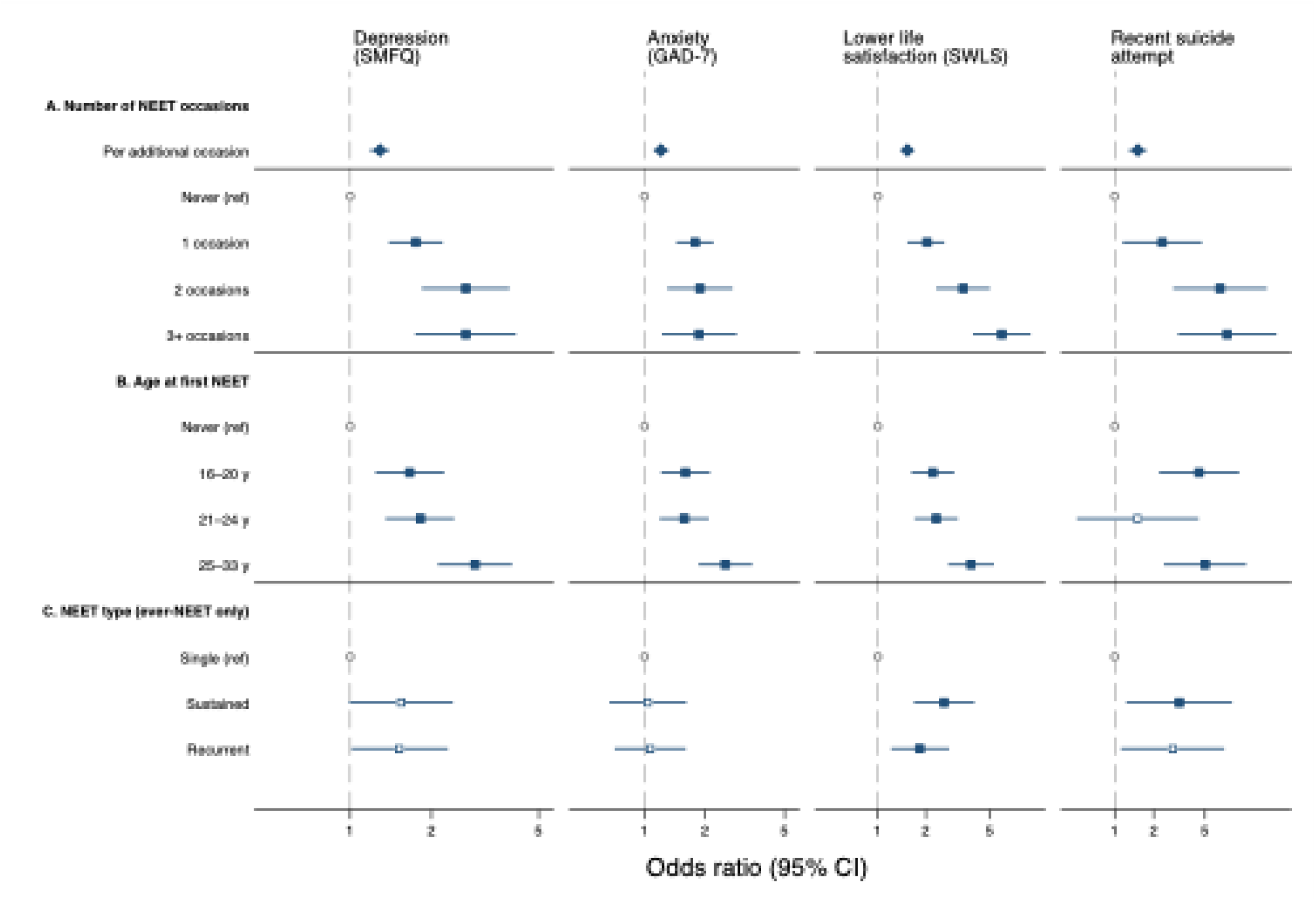
Association between NEET and mental health outcomes. SMFQ: Short Mood and Feelings Questionnaire; GAD-7: Generalised Anxiety Disorder-7; SWLS: Satisfaction with Life Survey. Solid shapes indicate result is significant after multiple correction (FDR).

Examining categories of NEET occasions, all four outcomes were elevated after a single occasion relative to never being NEET. Depression, lower life satisfaction and a recent suicide attempt increased further with additional occasions, whereas anxiety did not. The steepest gradients were observed for Depression (2.68 [1.74, 4.11], *P*_*FDR*_=1.76×10^-05^), lower life satisfaction (5.97 [3.93, 9.05], *P*_*FDR*_=4.94×10^-16^) and a recent suicide attempt (7.40 [3.08, 17.80], *P*_*FDR*_=1.18×10^-05^), all at three or more NEET occasions.

Associations between age of first NEET and mental health outcomes at age 33 were strongest for when age of first NEET occurred between the ages of 25 and 33 (Figure 1; Supplementary Table 2): depression (2.90 [2.11, 3.98], *P*_*FDR*_=2.98×10^-10^), anxiety (2.53 [1.86, 3.45], *P*_*FDR*_=1.42×10^-08^), lower life satisfaction (3.83 [2.76, 5.32], *P*_*FDR*_=1.15×10^-14^), suicide attempt (4.99 [2.40, 10.39], *P*_*FDR*_=3.50×10^-05^). In addition, first NEET between ages 16 and 20 was also associated with poorer mental health, most notably for suicide attempt following first NEET at ages 16 to 20 (4.49 [2.20, 9.17], *P*_*FDR*_=6.51×10^-05^), but also for depression (1.66 [1.24, 2.23], *P*_*FDR*_=0.0009), anxiety (1.60 [1.21, 2.12], *P*_*FDR*_=0.001), lower life satisfaction (2.21 [1.62, 3.01], *P*_*FDR*_=1.14×10^-06^). Results were broadly consistent when age of first NEET occurred between 21 and 24 years. Compared to those with a single occasion of NEET, sustained periods of NEET were associated with lower life satisfaction (2.60 [1.68, 4.02], *P*_*FDR*_=0.0001), and a recent suicide attempt (3.16 [1.23, 8.08], *P*_*FDR*_=0.042), and for life satisfaction with recurrent NEET occasions (1.83 [1.21, 2.80], *P*_*FDR*_=0.018),

Results were consistent with complete case analysis (Supplementary Tables 3 and 4).

## Discussion

Across a 17-year prospective period from adolescence to adulthood, accumulated NEET occasions were associated with depression, anxiety, lower life satisfaction and a recent suicide attempt at age 33. The magnitude of these associations differed by outcome. Individuals who had been NEET on three or more occasions were over 2 and half times more likely to report depression, approximately six times the odds of low life satisfaction and over seven times the odds of a recent suicide attempt, relative to those never NEET. Associations remained after adjustment for mental health at baseline.

These findings extend evidence from older cohorts. In the 1970 British Cohort study, psychological distress at age 51 rose steadily according to accumulated time spent in NEET between 16 and 24 years old [2]. Our results show the same graded pattern emerging by the early thirties in a cohort entering the labour market two decades later, and not solely investigating duration, but also onset and recurrence. Importantly, these results highlight that accumulated NEET (and not necessarily sustained or recurrent patterns) impacts beyond depression and anxiety into more severe outcomes such as a recent suicide attempt. Repeated disengagement may proliferate secondary stressors such as financial strain and delayed independence [12], which accumulate more in life satisfaction compared to depression and anxiety symptoms [10].

These patterns carry clinical and service level implications. Depression and anxiety were elevated after a single occasion, indicating mental health need should be identified at first disengagement, rather than reserved only for those disengaging repeatedly. The finding that a suicide attempt escalated with accumulation suggests preventing recurrence carries additional benefit, and that repeated NEET status may be a useful marker of suicide risk in services working with young adults. However, this runs counter to current UK Child and Adolescent Mental Health service architecture, since this support ceases for most at 18, precisely when NEET risk rises [5, 13].

An alternative explanation is selection. Poorer mental health could drive entry into and persistent NEET [14]. However, our estimates were adjusted for depression, anxiety and suicidal ideation measured at baseline, and for the strongest known antecedents of NEET, including childhood adversity, family socioeconomic position and academic attainment [15].

Associations were strongest for first NEET between ages 25 and 33. As this exposure is contemporaneous with outcome assessment, reverse causation is difficult to exclude for that age group. However, odds of a recent suicide attempt were comparably elevated following first NEET at ages 16 to 20, more than a decade before outcomes were assessed, suggesting risk is not solely a function of temporal proximity.

Strengths include the use of repeated prospective assessments of NEET over a 17-year window, and the ability to account for mental health information as baseline, mitigating some bias from reverse causation. Limitations include assessment at discrete waves, so occasions between assessments are unobserved and the count may conflate duration with recurrence and a regional cohort with limited ethnic diversity. This cohort also transitioned after the 2008 recession, when NEET experience was common and comparison groups were affected, so estimates may be conservative relative to cohorts transitioning in tighter labour markets.

## Data Availability

The informed consent obtained from ALSPAC (Avon Longitudinal Study of Parents and Children) participants does not allow the data to be made available through any third party maintained public repository. Supporting data are available from ALSPAC on request under the approved proposal number, B4792 and B5597. Full instructions for applying for data access can be found here: http://www.bristol.ac.uk/alspac/researchers/access/. The ALSPAC study website contains details of all available data (http://www.bristol.ac.uk/alspac/researchers/our-data/).

## Acknowledgments

We are extremely grateful to all the families who took part in this study, the midwives for their help in recruiting them, and the whole ALSPAC team, which includes data collection staff, data and administrations staff, technical managers and the technical staff with the Bristol Bioresource Laboratory, based within the University of Bristol.

## Contributions

Conceptualisation: Both authors. Funding acquisition: Both authors. Methodology: ASFK. Writing original draft: Both authors. Writing review and editing: Both authors.

