## Supplementary Information for "Not in Education Employment or Training (NEET) Trajectories Across Adolescence and Early Adulthood and Subsequent Mental Health"

**Further ALSPAC Sample Information**

Pregnant women resident in Avon, UK with expected dates of delivery between 1st April 1991 and 31st December 1992 were invited to take part in the study [1, 2]. 20,248 pregnancies have been identified as being eligible and the initial number of pregnancies enrolled was 14,541. Of the initial pregnancies, there was a total of 14,676 foetuses, resulting in 14,062 live births and 13,988 children who were alive at 1 year of age. When the oldest children were approximately 7 years of age, an attempt was made to bolster the initial sample with eligible cases who had failed to join the study originally. As a result, when considering variables collected from the age of seven onwards (and potentially abstracted from obstetric notes) there are data available for more than the 14,541 pregnancies mentioned above: The number of new pregnancies not in the initial sample (known as Phase I enrolment) that are currently represented in the released data and reflecting enrolment status at the age of 24 is 906, resulting in an additional 913 children being enrolled (456, 262 and 195 recruited during Phases II, III and IV respectively). The phases of enrolment are described in more detail in the cohort profile paper and its update. The total sample size for analyses using any data collected after the age of seven is therefore 15,447 pregnancies, resulting in 15,658 foetuses. Of these 14,901 children were alive at 1 year of age. From the age of 30, there were an additional 101 new participants recruited from 101 pregnancies during Phase V enrolment. The total sample size for analyses using any data collected after the age of 30 is therefore 15,548 pregnancies, resulting in 15,690 foetuses. Of these 15,002 children were alive at 1 year of age [3, 4].

Ethical approval for the study was obtained from the ALSPAC Ethics and Law Committee and the Local Research Ethics Committees. Informed consent for the use of all data collected was obtained from participants following the recommendations of the ALSPAC Ethics and Law Committee at the time. Participants can contact the study team at any time to retrospectively withdraw consent for their data to be used. Study participation is voluntary and during all data collection sweeps, information was provided on the intended use of data.

Please note that the study website contains details of all the data that is available through a fully searchable data dictionary and variable search tool: http://www.bristol.ac.uk/alspac/researchers/our-data/

Study data were collected and managed using REDCap electronic data capture tools hosted at the University of Bristol. REDCap (Research Electronic Data Capture) is a secure, web-based software platform designed to support data capture for research studies [5].

**Measures**

*Not in Education Employment and Training (NEET)*

NEET status was measured over 11 occasions at questionnaires CCS CCT CCU YPA YPB YPC YPE YPG YPJ YPP and the Focus@24 Clinic. We defined NEET status as individuals who reported currently being 1) employed and seeking work, being unable to work through sickness and/or disability, is currently doing voluntary work, or is currently out of work looking after family, at each questionnaire, respectively. We coded individuals at not NEET status if they reported any of the above, alongside being in full or part-time education and/or employment. We derived four measures of NEET from these variables:

First, a count of NEET occasions summed the number of assessments at which a participant met NEET criteria, ranging from 0 to 11. This variable was modelled continuously to estimate the association per additional occasion of NEET.

Second, a categorical measure of NEET occasions grouped participants into those never NEET, NEET on one occasion, NEET on two occasions, and NEET on three or more occasions. This allowed associations to be estimated without assuming a linear relationship between accumulated occasions and later outcomes.

Third, age at first NEET occasion classified participants according to the age at which they first met NEET criteria: never NEET, first NEET between ages 16 and 20 years, between 21 and 24 years, and between 25 and 33 years. Age bands were defined to correspond broadly to the transition out of compulsory education, early labour market entry, and established adulthood.

Finally, among participants ever NEET, the type of NEET distinguished those NEET on a single occasion only, those with sustained NEET (NEET at two or more consecutive assessments), or those with recurrent NEET (NEET at two or more non-consecutive assessments, indicating movement in and out of NEET status). Because this variable was defined only among those ever NEET, models used a single occasion as the reference category, isolating the contribution of exposure patterning from that of ever being NEET.

*Mental health outcomes (all assessed at YPP questionnaire)*

Depression was measured using the Short Mood and Feelings Questionnaire (SMFQ), a 13-item instrument examining depressive mood within the last two weeks [6]. The SMFQ scores range between 0-26 with higher scores indicting higher depressive symptoms. Scores of ≥11 on the SMFQ have good specificity and sensitivity for probable depression [7, 8]. We coded those with ≥11 as having depression for this analysis.

Anxiety was measured using the Generalised Anxiety Disorder Assessment (GAD-7), a 7-item instrument which measures the presence of generalised anxiety disorder symptoms within the last two weeks [9]. The GAD-7 scores range between 0-21 with higher scores indicting higher anxiety symptoms and scores ≥10 have good specificity and sensitivity for probable generalised anxiety disorder [10]. We coded those with ≥10 as having anxiety for this analysis.

Life satisfaction was measured using the Satisfaction with Life Scale (SWLS), a 5-item instrument which measures overall life satisfaction [11]. The SWLS scores between 7 and 35, with higher scores indicating higher life satisfaction. We coded those with ≤14 as having lower life satisfaction for this analysis.

A recent suicide attempt was measured using a single item which asked: “Since the start of 2023, have you attempted suicide, and if so, how did this affect you?”. We coded individuals who answered yes, regardless of whether it affected them as having a recent suicide attempt in this analysis.

**Multiple imputation**

Missing covariate data were addressed using multiple imputation by chained equations (MICE) in Stata 18, implemented with the mi impute chained command.

Exposure (derived NEET variables) and outcome variables (mental health outcomes) were fully observed and not imputed. NEET measures (occasion count, categorical occasions, age at first NEET, NEET type, all age 33 outcomes (depression, anxiety, past-year suicide attempt, life satisfaction, risky alcohol use, weekly smoking), the number of completed NEET assessments and sex were treated as complete and registered as regular variables. The analytic sample was therefore restricted to participants with observed exposure and outcome data (n=3792), with imputation used to recover missing covariate information within this sample.

Covariates were imputed based using models appropriate to their measurement scale. Binary variables — ethnicity, maternal social class at birth, anxiety at age 17, and baseline suicidal ideation, risky alcohol use, smoking, maternal smoking during pregnancy, and repeated adolescent depressive symptom (SMFQ) measures between 11 and 18 were imputed by logistic regression with the augmented approach to accommodate perfect prediction. Ordinal variables (maternal education at birth, number of childhood traumas between 0-17 years, parity) were imputed by ordered logistic regression, also augmented. Continuous variables: maternal postnatal depression at three occasions (EPDS), academic attainment at Key Stages 1 to 4, three repeated measures of emotional symptoms, conduct problems (both SDQ) and three repeated measures of financial problems were imputed by predictive mean matching drawing from the 10 nearest neighbours, which avoids imposing normality on skewed distributions. The standardised polygenic score for major depressive disorder (taken from the latest GWAS of depression) was imputed by linear regression.

All imputation models included the exposure, outcome and complete variables listed above as predictors, ensuring congeniality with the substantive analysis models. Fifty imputed datasets were generated, a number chosen to exceed the proportion of incomplete cases and to yield stable standard errors. A fixed random seed (20260831) was set for reproducibility. Convergence was assessed by inspecting trace plots of the imputed means and standard deviations across iterations for each imputed variable. Estimates were combined across datasets using Rubin's rules.

| **Supplementary Table 1. Association between NEET occasions between 16-33 years and mental health outcomes at 33 years using imputed data (n=3792)** | | | | |
| --- | --- | --- | --- | --- |
|  | Depression  (SMFQ)^a^ | Anxiety  (GAD-7)^b^ | Lower Life Satisfaction (SWLS)^a^ | Recent Suicide  Attempt^c^ |
|  | OR (95% CI),  *P_FDR_* | OR (95% CI),  *P_FDR_* | OR (95% CI),  *P_FDR_* | OR (95% CI),  *P_FDR_* |
| Per NEET occasion (count) | 1.29 (1.18, 1.40),  *P*=3.04x10^-08^ | 1.21 (1.11, 1.32),  *P*=7.54 x10^-05^ | 1.53 (1.40, 1.68),  *P*=2.26 x10^-19^ | 1.51 (1.28, 1.77),  *P*=1.17 x10^-06^ |
| NEET category  (ref=0 vs.) |  |  |  |  |
| 1 occasion | 1.75 (1.39, 2.20), *P*=3.01x10^-06^ | 1.79 (1.44, 2.21), *P*=3.63x10-^07^ | 2.03 (1.53, 2.61),  *P*=1.08x10^-07^ | 2.33 (1.15, 4.72),  *P*=0.019 |
| 2 occasions | 2.68 (1.84, 3.89), *P*=5.45x10^-07^ | 1.89 (1.30, 2.75),  *P*=0.002 | 3.42 (2.32, 5.04),  *P*=3.67x10^-09^ | 6.52 (2.83, 15.01), *P*=1.40x10^-05^ |
| 3+ occasions | 2.68 (1.74, 4.11), *P*=1.76x10^-05^ | 1.87 (1.22, 2.88),  *P*=0.004 | 5.97 (3.93, 9.05),  *P*=4.94x10^-16^ | 7.40 (3.08, 17.80), *P*=1.18x10^-05^ |
| Imputed data analysis, adjusted for sex, ethnicity, maternal education at birth, maternal social class at birth, maternal financial problems at birth, maternal postnatal depression, number of childhood traumas, childhood educational attainment (key stage 2 scores) and the number of completed NEET questionnaires. OR: Odds Ratio; CI: Confidence Interval; *P_FDR_*: P-value corrected for Benjamini-Hochberg false discovery rate. SMFQ: Short Mood and Feelings Questionnaire; GAD-7: Generalised Anxiety Disorder-7; SWLS: Satisfaction with Life Survey.   ^a^ adjusted for depression at baseline/age 16 (SMFQ) ^b^ adjusted for anxiety at age 17 (Clinical Interview Schedule-Revised) ^a^ adjusted for depression at baseline/age 16 (SMFQ) ^c^ adjusted for suicidal ideation at baseline/age 16 | | | | |

| **Supplementary Table 2. Association between age of first NEET and NEET type between 16-33 years and mental health outcomes at 33 years using imputed data (n=3792)** | | | | |
| --- | --- | --- | --- | --- |
|  | Depression  (SMFQ)^a^ | Anxiety  (GAD-7)^b^ | Lower Life Satisfaction (SWLS)^a^ | Recent Suicide Attempt^c^ |
|  | OR (95% CI),  *P_FDR_* | OR (95% CI),  *P_FDR_* | OR (95% CI),  *P_FDR_* | OR (95% CI),  *P_FDR_* |
| NEET onset (ref=never vs.) |  |  |  |  |
| First NEET  (ages 16-20) | 1.66 (1.24, 2.23),  *P*=0.0009 | 1.60 (1.21, 2.12),  *P*=0.001 | 2.21 (1.62, 3.01),  *P*=1.14x10^-06^ | 4.49 (2.20, 9.17),  *P*=6.51 x10^-05^ |
| First NEET  (ages 21-24) | 1.82 (1.35, 2.43), *P=*9.90x10^-05^ | 1.58 (1.19, 2.09),  *P=*0.002 | 2.32 (1.70, 3.17),  *P=*3.12x10^-07^ | 1.50 (0.51, 4.45), *P=*0.464 |
| First NEET  (ages 25-33) | 2.90 (2.11, 3.98), *P*=2.98x10^-10^ | 2.53 (1.86, 3.45), *P*=1.42x10^-08^ | 3.83 (2.76, 5.32),  *P*=1.15x10^-14^ | 4.99 (2.40, 10.39),  *P*=3.50x10^-05^ |
| NEET type  (ref=Single occasion vs.) |  |  |  |  |
| Single sustained occasion  (2+ occasions) | 1.54 (0.99, 2.40),  *P=*0.073 | 1.04 (0.67, 1.62),  *P=*0.868 | 2.60 (1.68, 4.02),  *P=*0.0001 | 3.16 (1.23, 8.08),  *P=*0.042 |
| Recurrent occasions (in and out) | 1.52 (1.01, 2.30),  *P=*0.073 | 1.07 (0.71, 1.61),  *P=*0.852 | 1.83 (1.21, 2.80),  *P=*0.018 | 2.81 (1.12, 7.04),  *P=*0.054 |
| Imputed data analysis, adjusted for sex, ethnicity, maternal education at birth, maternal social class at birth, maternal financial problems at birth, maternal postnatal depression, number of childhood traumas, childhood educational attainment (key stage 2 scores) and the number of completed NEET questionnaires. OR: Odds Ratio; CI: Confidence Interval; *P_FDR_*: P-value corrected for Benjamini-Hochberg false discovery rate. SMFQ: Short Mood and Feelings Questionnaire; GAD-7: Generalised Anxiety Disorder-7; SWLS: Satisfaction with Life Survey. ^a^ adjusted for depression at baseline (SMFQ) ^b^ adjusted for anxiety at age 17 (CISR) ^a^ adjusted for depression at baseline (SMFQ) ^c^ adjusted for suicidal ideation at baseline | | | | |

| **Supplementary Table 3. Association between NEET occasions between 16-33 years and mental health outcomes at 33 years using complete data** | | | | |
| --- | --- | --- | --- | --- |
|  | Depression  (SMFQ)^a^ | Anxiety  (GAD-7)^b^ | Lower Life Satisfaction (SWLS)^a^ | Recent Suicide  Attempt^c^ |
|  | OR (95% CI),  *P* (n=1758) | OR (95% CI),  *P* (n=1619) | OR (95% CI),  *P* (n=1758) | OR (95% CI),  *P* (n=1791) |
| Per NEET occasion (count) | 1.32 (1.16, 1.51),  *P*=0.00003 | 1.22 (1.06, 1.42),  *P*=0.006 | 1.45 (1.27, 1.66),  *P*=3.79 x10^-08^ | 1.53 (1.19, 1.96),  *P*=0.0009 |
| NEET category  (ref=0 vs.) |  |  |  |  |
| 1 occasion | 1.75 (1.25, 2.44),  *P*=0.001 | 1.59 (1.39, 2.23), *P*=0.007 | 1.64 (1.12, 2.39),  *P*=0.011 | 2.13 (0.71, 6.34),  *P*=0.175 |
| 2 occasions | 2.55 (1.51, 4.29),  *P*=0.0005 | 1.77 (1.00, 3.12),  *P*=0.049 | 2.74 (1.57, 2.39),  *P*=0.0004 | 3.47 (0.71, 13.39),  *P*=0.071 |
| 3+ occasions | 3.18 (1.65, 6.11), *P*=0.0005 | 1.89 (0.89, 3.99),  *P*=0.095 | 6.64 (3.51, 12.57),  *P*=5.91x10^-09^ | 7.53 (2.05, 27.68), *P*=0.002 |
| Complete case analysis, adjusted for sex, ethnicity, maternal education at birth, maternal social class at birth, maternal financial problems at birth, maternal postnatal depression, number of childhood traumas, childhood educational attainment (key stage 2 scores) and the number of completed NEET questionnaires. OR: Odds Ratio; CI: Confidence Interval; SMFQ: Short Mood and Feelings Questionnaire; GAD-7: Generalised Anxiety Disorder-7; SWLS: Satisfaction with Life Survey,  ^a^ adjusted for depression at baseline/age 16 (SMFQ) ^b^ adjusted for anxiety at age 17 (Clinical Interview Schedule-Revised) ^a^ adjusted for depression at baseline/age 16 (SMFQ) ^c^ adjusted for suicidal ideation at baseline/age 16 | | | | |

| **Supplementary Table 4. Association between age of first NEET and NEET type between 16-33 years and mental health outcomes at 33 years using complete case data** | | | | |
| --- | --- | --- | --- | --- |
|  | Depression  (SMFQ)^a^ | Anxiety  (GAD-7)^b^ | Lower Life Satisfaction (SWLS)^a^ | Recent Suicide  Attempt^c^ |
|  | OR (95% CI),  *P* (n=1758) | OR (95% CI),  *P* (n=1619) | OR (95% CI),  *P* (n=1758) | OR (95% CI),  *P* (n=1791) |
| NEET onset (ref=never vs.) |  |  |  |  |
| First NEET  (ages 16-20) | 1.76 (1.18, 2.64),  *P*=0.006 | 1.44 (0.93, 2.32),  *P*=0.102 | 2.20 (1.44, 3.36),  *P*=0.0003 | 4.06 (1.45, 11.33),  *P*=0.008 |
| First NEET  (ages 21-24) | 1.72 (1.14, 2.62),  *P=*0.010 | 1.32 (0.85, 2.05),  *P=*0.216 | 1.79 (1.13, 2.85),  *P=*0.013 | 1.34 (0.29, 6.31), *P=*0.707 |
| First NEET  (ages 25-33) | 3.57 (2.15, 5.92), *P*=8.50x10^-07^ | 2.93 (1.78, 4.84), *P*=0.00003 | 3.31 (1.92, 5.71),  *P*=0.00002 | 4.51 (1.20, 16.95),  *P*=0.026 |
| NEET type  (ref=Single occasion vs.) |  |  |  |  |
| Single sustained occasion  (2+ occasions) | 1.65 (0.84, 3.25),  *P=*0.146 | 1.22 (0.56, 2.68),  *P=*0.610 | 2.36 (1.18, 4.72),  *P=*0.015 | 2.55 (0.55, 11.81),  *P=*0.230 |
| Recurrent occasions (in and out) | 1.54 (0.87, 2.75),  *P=*0.140 | 1.09 (0.59, 2.02),  *P=*0.776 | 2.38 (1.32, 4.30),  *P=*0.004 | 2.20 (0.56, 8.65),  *P=*0.259 |
| Complete case analysis, adjusted for sex, ethnicity, maternal education at birth, maternal social class at birth, maternal financial problems at birth, maternal postnatal depression, number of childhood traumas, childhood educational attainment (key stage 2 scores) and the number of completed NEET questionnaires. OR: Odds Ratio; CI: Confidence Interval; SMFQ: Short Mood and Feelings Questionnaire; GAD-7: Generalised Anxiety Disorder-7; SWLS: Satisfaction with Life Survey. ^a^ adjusted for depression at baseline (SMFQ) ^b^ adjusted for anxiety at age 17 (CISR) ^a^ adjusted for depression at baseline (SMFQ) ^c^ adjusted for suicidal ideation at baseline | | | | |
